# Is point wise analysis of the Humphrey visual field feasible as a primary outcome in idiopathic intracranial hypertension?

**DOI:** 10.1101/2022.05.30.22275503

**Authors:** Susan P Mollan, Samuel Bodoza, Áine Ní Mhéalóid, James L Mitchell, Neil R. Miller, Giovanni Montesano, David P Crabb, Michael Wall, Kristian Brock, Alexandra J Sinclair

## Abstract

**Purpose:** Using the Idiopathic Intracranial Hypertension Weight Trial (IIH:WT) data, this study aimed to determine if point analysis of the Humphrey visual field (HVF) could be more informative than the perimetric mean deviation (PMD) as an IIH trial outcome measure.

**Methods:** IIH:WT was a randomized controlled trial that recruited 66 people with active IIH (mean ± standard deviation age 32 ± 7.8 years). Event-based analysis using a pointwise analysis of the numerical sensitivity data was performed. The number of participants that would be eligible for analysis was calculated when the data were enriched to reflect a medically treated cohort defined as a PMD between -2dB to -7dB.

**Results:** The HVF 24-2 mean ± standard deviation PMD in the worse eye was ™3.5 ± 1.1dB, (range,™2.0 to ™6.4dB), and point sensitivity showed a preference for peripheral and blind spot locations. Those points between 0 and -10dB demonstrated negligible ability to improve compared with those between -10dB and -25dB. In evaluating feasibility for a medical intervention trial, 346 points were available for analysis between -10dB and -25dB bilaterally compared with 4123 in baseline sensitivities of 0 to -10dB.

**Conclusions:** Mildly affected baseline sensitivities were unlikely to show considerable change over 24 months. There were fewer points available for analysis and greater variability if moderately affected baseline points were chosen. If point analysis was used as an outcome measure in a medical intervention trial, the majority of points would not demonstrate clinically meaningful change, thus offering little advantage over PMD.

## Introduction

Idiopathic Intracranial Hypertension (IIH) is characterised by raised intracranial pressure (ICP) that in most cases causes papilloedema with risk of permanent visual loss.^1^ It is associated with systemic metabolic dysfunction and central obesity.^2,3,4^ The incidence of IIH is increasing in line with the escalation in worldwide obesity rates.^5,6^ The IIH weight trial (IIH:WT) provided evidence that weight loss through bariatric surgery resulted in long-term remission of ICP compared with a lifestyle weight management intervention.7

Measurable outcomes are vital to provide evidence when treating patients,^8^ and the perimetric mean deviation (PMD), among other end points such as lumbar puncture opening pressure, has been used as an end point in IIH clinical trials.9 There are recognised limitations of visual field testing in IIH, such as the time needed to complete the test and the need to perform accurate reliable visual field testing.^10^ There is thus an established performance learning effect that impacts accuracy of initial performances.^11^ It has also been recently been observed that cognitive dysfunction that occurs in patients with IIH can affect the reliability of visual fields.^12^ In the IIH Treatment Trial (IIHTT), a 0.71dB difference in the PMD was found between the two trial arms and was related to clinically significant findings, such as noteworthy improvements in papilloedema, ICP as measured by lumbar puncture, and improved quality-of-life measures.^9,13^

There are a number of different ways to evaluate of visual field damage.^14,15,16,17^ PMD, is an event based analysis, as implemented in the Humphrey Field Analyzer (HFA, Zeiss Meditec, Dublin, CA), is measured in decibels (dB) using a logarithmic scale and is calculated from all of the total deviation points with weighting inversely proportional to the expected variance at each location in a normal population, effectively giving more weight to the central locations.^18-20^ However, peripheral locations are more affected than central locations in patients with IIH.^17^ Pointwise approaches have the advantage of being more sensitive to localized loss than other types of global analyses as they retain the spatial representation of the visual field. Pointwise analysis of the IIHTT detailed significant improvement in the visual function in the active treatment arm compared with placebo.^14^ To apply this analysis to future trials, or indeed to consider restricting the analysis to a particular subset of points in the visual field, the number and location of points that could be predicted to change in an IIH intervention trial needs to be determined. In addition, the amount of change in these points needs to be established to determine if sufficient points have the ability to change to an extent that would be clinically meaningful. The purpose of this study was to characterise the pointwise pattern of visual field change in a cohort of people with active IIH who were in a setting of a randomised clinical trial and to determine if point analysis would be feasible as an outcome measure for future IIH clinical trials.

## Materials and methods

IIH:WT was a prospective, multi-center, open-label, parallel-group, controlled trial in which participants with IIH were randomized in a 1:1 ratio to a bariatric surgery pathway or the Weight Watchers™ programme, a community weight management intervention (CWI). The study was approved by the Ethics Review Board of the National Research Ethics Committee West Midlands –The Black Country approved IIH:WT (14/WM/0011). In accordance with the Declaration of Helsinki, all subjects gave written informed consent to participate in the study, and the detailed clinical trial methodology has been published.^21^

### Subjects

Women (18-55 years) with a body mass index over 35 kg/m^2^ were eligible if they had a clinical diagnosis of active IIH meeting the criteria outlined by Friedman et al.^22^ All participants were recruited between March 2014 and October 2017.

Evaluations were performed at baseline, 12 months, and 24 months.^21^ The primary outcome was ICP as measured by lumbar puncture; secondary outcomes included lumbar puncture opening pressure at 24 months as well as visual acuity, contrast sensitivity, PMD, and quality-of-life measures. Optic nerve head swelling was measured using spectral domain optical coherence tomography (SD-OCT; Spectralis, Heidelberg Engineering). Three masked neuro-ophthalmologists used colour fundus photographs to assess the severity of papilloedema using the Frisén classification grades of 0 to 5.^23^

At each visit, Humphrey Visual Fields with a 24-2 Swedish Interactive Threshold Algorithm (SITA) standard test pattern using a size III white stimulus were performed. HVF were included for analysis if they were considered reliable as defined by less than 15% false-positive rates and 30% fixation losses and false-negative rates according to previous criteria.^24^ In this analysis, the raw values of the patient’s retinal sensitivity at each of the HVF 24-2 predetermined points were extracted from pdf scans of the HVFs using a custom data extraction tool based around the Python^25^ package ‘hvf extraction script’.^26^ This script used Google’s Tesseract Optical Character Recognition^27^ to distinguish text inside a digital image and return the relevant text into a useable format. Although the ‘hvf extraction script’ was not originally intended for use on scanned documents, we found that by cleaning the images before processing, we could obtain values for a majority of the retinal sensitivity points. To account for any missing data between the original values and the data extraction tool, we completed a manual validation of the whole cohort point retinal sensitivity to eliminate missing data and to detect any discrepancies.

### Statistical Analysis

Analysis of clinical data was based on the full data set according to the statistical analysis plan.7 In this evaluation, analyses were based on a per protocol analysis. Statistical analysis was performed using R v3·6·3 (R Foundation for Statistical Computing, Vienna, Austria). Data were reported with mean and standard deviations (SD) for normally distributed variables (median and range for non-parametric data). Hierarchical linear regression models were used to analyse repeated measures of the primary and secondary outcomes and to estimate differences adjusted for baseline values. In these models, population-level effects (also known as fixed effects) comprised the intercept, time as a factor variable, and the 2-way interaction of treatment arm and time as a factor variable to model-changing treatment effects over time. Group-level effects (also known as random effects) comprised patient-level adjustments to the intercept. Missing clinical data, due to any absence or choice, were excluded from the analysis and not imputed. The threshold for statistical significance was pre-specified at p<0.05.

## Results

Characteristics of the study population are summarized in Table 1. Headache was the predominant reported symptom and also was the most vexing symptom when participants ranked the importance of their symptoms; visual symptoms, as defined by the participants, were reported by 77% (supplemental table 1) and were the second-most bothersome aspect of the condition.

**Table 1.**
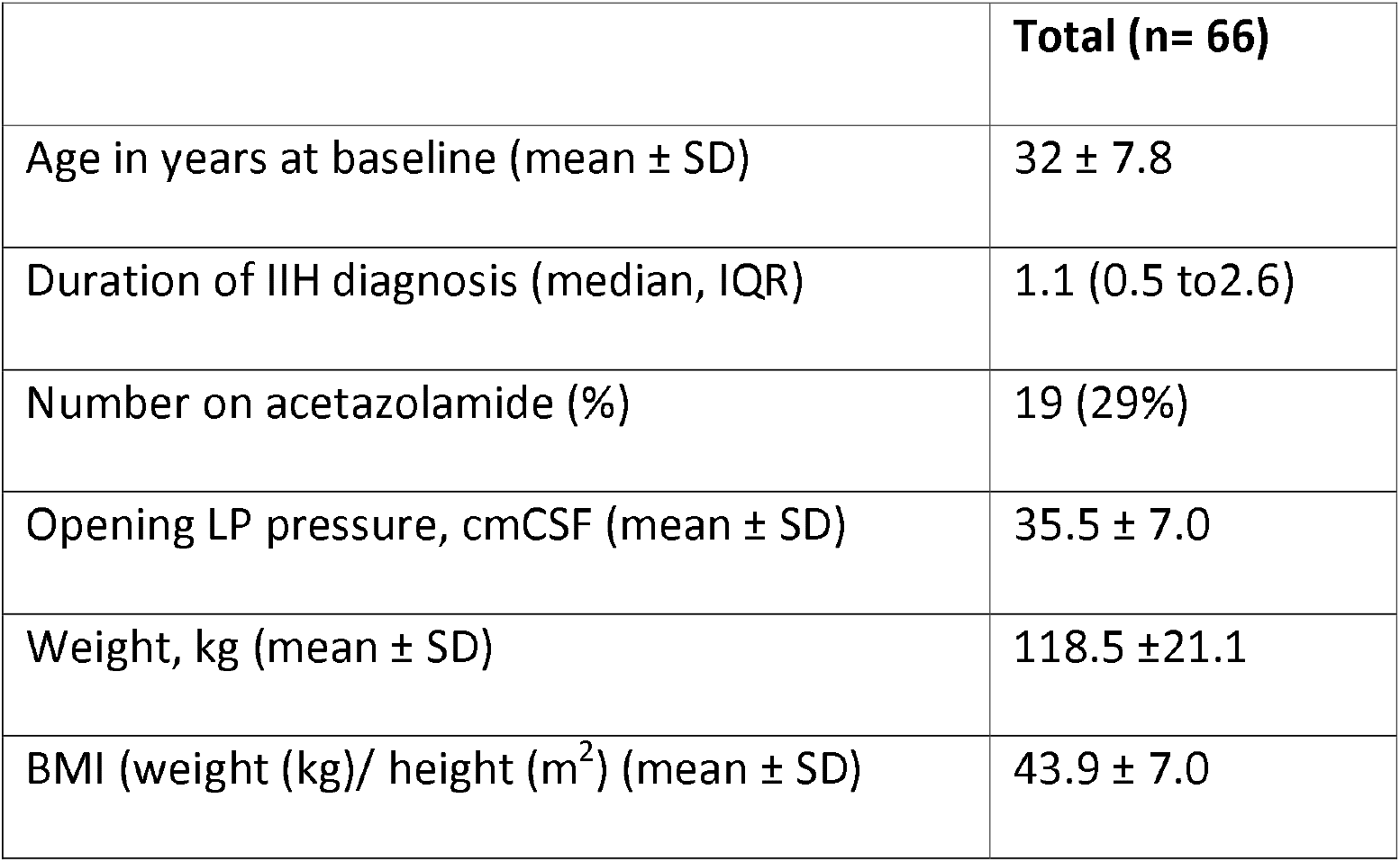
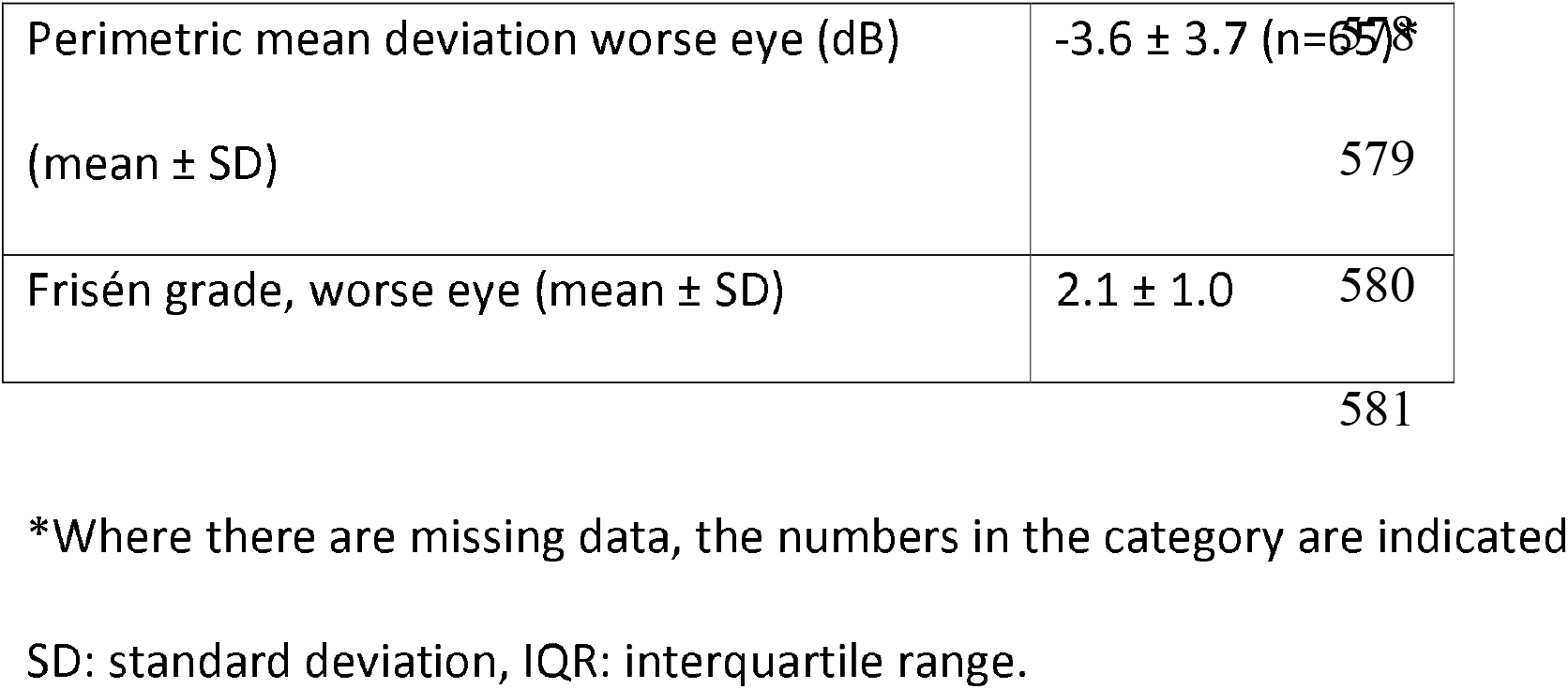
IIH:WT baseline characteristics.

The cohort had marked variability in PMD (worse eye) of -3.6 ± 3.7 dB (mean ± SD) (n=65) at baseline; -2.4 ± 2.5dB (n= 58) at 12 months; and -3.1 ± 2.5 dB at 24 months. There was improvement in the PMD in both trial arms. In the bariatric surgery arm, the PMD improved from -3.6 ± 3.8 dB to -2.8 ± 2.6 dB at 12 months, and in the CWI arm, from -3.5 ± 3.8 dB to - 2.0 ± 2.3 dB (adjusted mean difference= -0.5, 95% Confidence Interval (CI)): -2.0 - 1.0, p=0.526) (Table 2). Papilloedema decreased in both arms, with a median Frisén grade of 2 (IQR 2-3) at baseline, decreasing to 1 (IQR 1-2) in the bariatric arm, and median Frisén grade 2 (IQR 2-3) decreasing to 1 (IQR 1-2) in the CWI arm at 12 months. This was reflected in the global RNFL OCT imaging, showing significant improvement in both arms (Table 2). These improvements occurred in the context of lowering ICP over the course of the study, with the mean ± SD LP opening pressure decreasing from 34.8 ± 5.8 cm CSF at baseline to 26.4 ± 8.1 cm CSF at 12 months in the bariatric surgery arm (−8.7 ± 1.3, p<0.001). In the CWI arm, the mean ± SD LP opening pressure decreased from 34.6 ± 5.6 cm CSF at baseline to 32.0 ± 5.2 cm CSF at 12 months (−2.5 ± 1.4, p=0.084). The adjusted mean difference between the two arms was -6.0 ± 1.8, p=0.001 at 12 months and -8.2 ± 2.0, p<0.001 at 24 months.^7^

**Table 2.** Visual function and optic nerve head status as measured by SD-OCT at baseline and 12 months

|  | Baseline<br>mean value (±SD),<br>number |  | 12 months<br>mean value (±SD),<br>number |  | Adjusted mean<br>difference in change<br>at 12 months (95%<br>C.I), p |  | Difference<br>between arms<br>at 12 months |
| --- | --- | --- | --- | --- | --- | --- | --- |
|  | Bariatric<br>surgery | CWI | Bariatric<br>surgery | CWI | Bariatric<br>surgery | CWI | Hierarchical<br>regression<br>mean (SE);<br>95%CI, p |
| **Visual**<br>**Acuity**<br>**LogMAR** | 0.0<br>(0.2), 33 | 0.0<br>(0.2),<br>33 | 0.0 (0.2),<br>30 | 0.0<br>(0.2),<br>28 | -0.1 (0.1);<br>(-0.2, 0.0),<br>p=0.058 | 0.0 (0.1);<br>(-0.1,<br>0.1),<br>p=0.985 | 0.0 (0.1); (-0.1,<br>0.1), p=0.598 |
| **Contrast**<br>**Sensitivity** | 1.7<br>(0.1), 33 | 1.7<br>(0.1),<br>33 | 1.7 (0.1),<br>29 | 1.7<br>(0.1),<br>28 | 0.0 (0.1);<br>(0.0, 0.1),<br>p=0.463 | 0.0 (0.1);<br>(0.0,<br>0.1),<br>p=0.630 | 0.0 (0.1); (-0.1,<br>0.0), p=0.411 |
| **Perimet**<br>**ric**<br>**Mean**<br>**Deviation** | -3.6<br>(3.5), 32 | -3.5<br>(3.8),<br>33 | -2.8<br>(2.6), 29 | -2.0<br>(2.3),<br>29 | 1.0 (0.5);<br>(-0.1, 2.0),<br>p=0.064 | 1.3 (0.5);<br>( 0.3,<br>2.3),<br>p=0.010 | -0.5 (0.8); (-<br>2.0, 1.0),<br>p=0.526 |
| **OCT**<br>**RNFL**<br>**Average** | 148.8<br>(99.1),<br>32 | 161.7<br>(95.7),<br>32 | 103.0<br>(27.4),<br>29 | 111.8<br>(33.1),<br>28 | -45.3<br>(15.7); (-<br>76.1, - | -50.5<br>(15.9); (-<br>81.7, - | -8.1 (17.3); (-<br>41.9, 25.8),<br>p=0.641 |
| ($\mu\text{m}$) | | | | | 14.5),<br>p=0.004 | 19.3),<br>p=0.001 | |

All measures are of worse eye. Negative values in the mean difference and adjusted mean difference favour surgical arm. C.I: confidence interval; SD-OCT: spectral-domain optical coherence tomography; RNFL: retinal nerve fibre layer; SD: standard deviation.

### Pointwise sensitivity – whole cohort at baseline

The whole cohort HVF were characterised at baseline by the sensitivity of each point on the 24-2. This showed that the central points were less affected than the peripheral points (Figure 1a). The whole cohort then was categorised according to the extent of their reduced visual function at baseline as per PMD category of ≥ -2dB (Figure 1b); between -2dB to -7Db (Figure 1c); and ≤-7dB (Figure 1d). As the visual function declined, the distribution of the average deviation points becomes increasing prominent in the periphery and around the blind spot.

**Figure 1.**
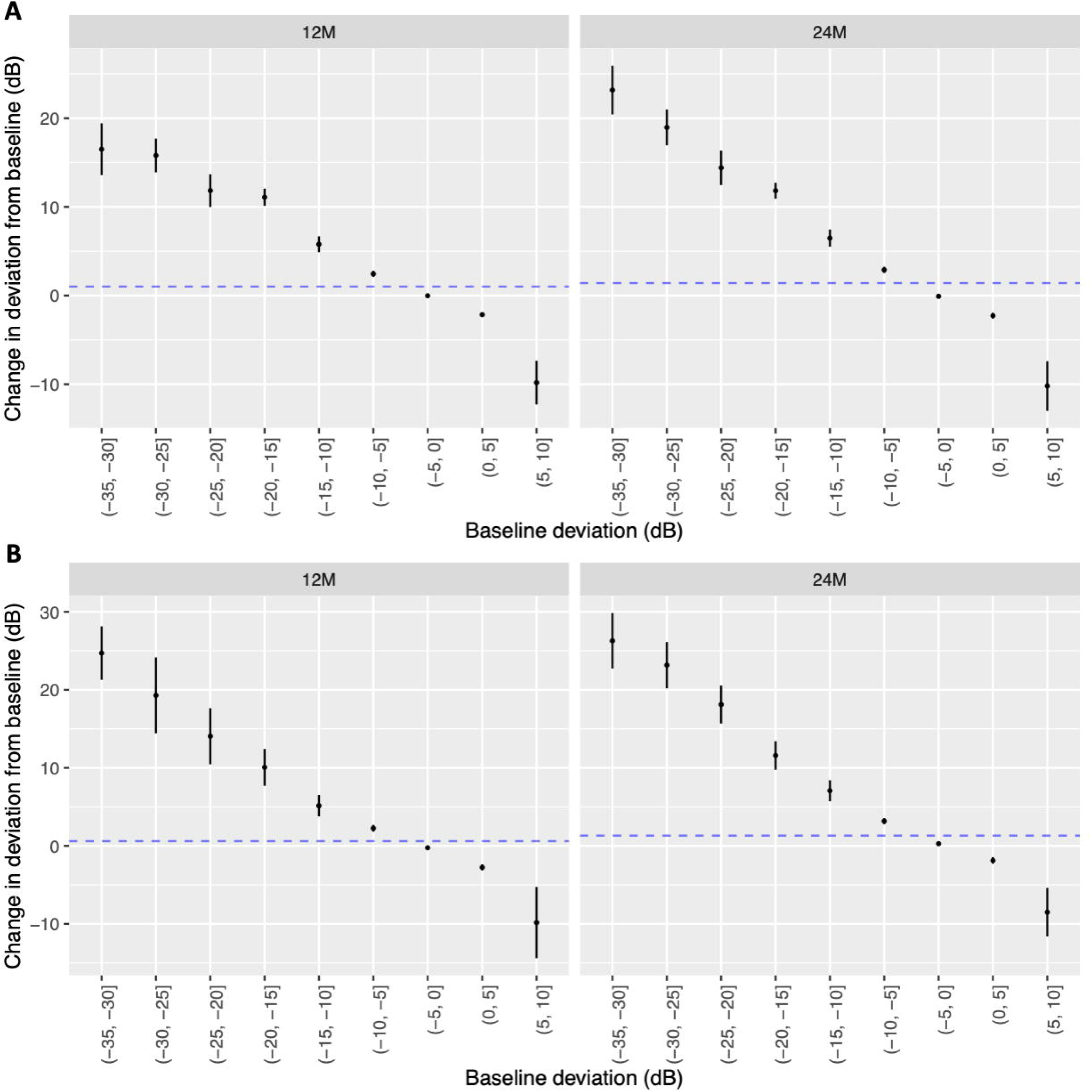
A) The retinal sensitivity at baseline for each location of the Humphrey visual field for the whole cohort, within eye. All patients are used. B) The retinal sensitivity at baseline for each location of the visual field for those with a PMD of -2dB or better. C) The retinal sensitivity at baseline for each location of the visual field for those with a PMD between -2dB and -7dB. D) The retinal sensitivity at baseline for each location of the visual field for those with a PMD worse than -7dB.

### Pointwise location sensitivity – whole cohort at 12 and 24 months

To understand the pointwise change over the course of the study, we categorised the points by their individual pointwise sensitivity at baseline. We then plotted the mean change in sensitivity at each point from the visual field from baseline to 12 and 24 months (Figure 2). Points with baseline sensitivities between 0 and -10dB showed small changes over time points (0 to 5dB, mean 0.02 ± 3.1; -5 to -10dB, mean 2.4 ± 4.7 at 12 months) (Figure 2; Table 3). Points with sensitivities worse than -10dB demonstrated a larger improvement over time; for example, between -10 and -15dB, mean ± SD was 5.78 ± 6.10dB; -15 to -20dB, mean was 11.10 ± 5.02dB (Figure 2; Table 3) at 12 months. For points with baseline sensitivities of -35 to -30dB, there was a large standard deviation (mean, 16.51 ± 13.75dB) (Figure 2; Table 3) at 12 months.

**Figure 2.**
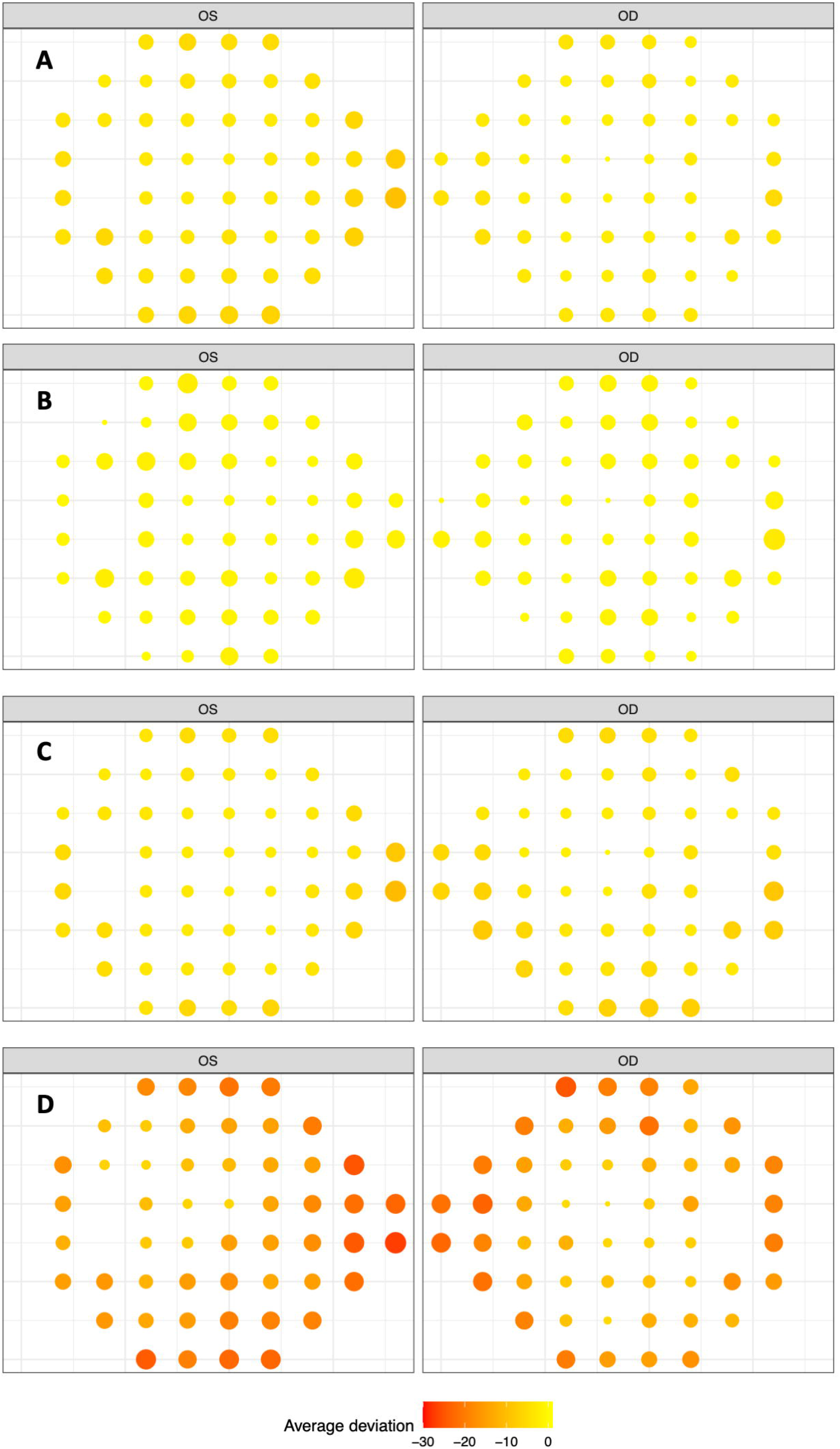
A) The mean change in deviation from baseline (and 95% confidence intervals) to 12 months and 24 months in subsets of points classified by baseline deviation. All patient eyes are used. Categories with at least 10 observations at each time point are shown. The sizes of the groups are naturally different, and this is reflected in the widths of the confidence intervals. B) The mean change in deviation from baseline (and 95% confidence intervals) to 12 months and 24 months in the population defined by a PMD between -2dB and -7dB at baseline (simulation of a medically treated population).

**Table 3.** Number of points and mean change in point sensitivity in test locations categorized by the baseline point sensitivity subgroup at 12 and 24 months.

| Time (months) | Baseline point sensitivity subgroup (dB) | Number of points | Mean at baseline (dB) | Mean change from baseline | Standard deviation |
| --- | --- | --- | --- | --- | --- |
| 12 | [-35, -30] | 85 | -32.211765 | 16.5058824 | 13.745669 |
|  | [-30, -25] | 56 | -26.75 | 15.8035714 | 7.24987685 |
|  | [-25, -20] | 67 | -21.940299 | 11.8358209 | 7.71970524 |
|  | [-20, -15] | 103 | -17.029126 | 11.0873786 | 5.01977813 |
|  | [-15, -10] | 176 | -11.721591 | 5.78409091 | 6.07843751 |
|  | [-10, -5] | 711 | -6.3614627 | 2.44303797 | 4.74140335 |
|  | [-5, 0] | 3412 | -1.6770223 | -0.0219812 | 3.03103034 |
|  | [0, 5] | 661 | 1.56278366 | -2.1527988 | 3.10219065 |
|  | [5, 10] | 11 | 7.45454545 | -9.8181818 | 4.16696969 |
| 24 | [-35, -30] | 63 | -31.984127 | 23.1746032 | 11.1406545 |
|  | [-30, -25] | 53 | -26.773585 | 18.9622642 | 7.5242231 |
|  | [-25, -20] | 61 | -21.868852 | 14.4098361 | 7.73816742 |
|  | [-20, -15] | 102 | -17.009804 | 11.8235294 | 4.64462164 |
|  | [-15, -10] | 166 | -11.73494 | 6.47590361 | 6.37200488 |
|  | [-10, -5] | 652 | -6.4033742 | 2.89723926 | 4.54979533 |
|  | [-5, 0] | 2784 | -1.7406609 | -0.0858477 | 3.78872898 |
|  | [0, 5] | 480 | 1.65 | -2.2729167 | 3.66420589 |
|  | [5, 10] | 10 | 7.4 | -10.2 | 4.51663592 |

When points with baseline sensitivity between -10 and -25dB were analysed for the whole cohort, mean change at 12 months was 8.53 ± 6.75dB, increasing further to 9.61 ± 6.99dB by 24 months. (Table 3; Table 4)

**Table 4.** Number of points and mean change in point sensitivity over time in test locations with a baseline point sensitivity between -10 and -25dB, categorized by trial arm and use of acetazolamide.

| Time (months) | Group | Number of points between -10 and -25dB | Mean | Standard deviation |
| --- | --- | --- | --- | --- |
| 12 | Whole cohort | 346 | 8.53 | 6.75 |
|  | CWI | 127 | 8.16 | 7.15 |
|  | CWI, Acetazolamide | 22 | 8.77 | 4.31 |
|  | CWI, No acetazolamide | 105 | 8.03 | 7.62 |
|  | Bariatric surgery | 219 | 8.75 | 6.51 |
| 24 | Whole cohort | 329 | 9.60 | 6.99 |
|  | CWI | 118 | 11.50 | 7.09 |
|  | CWI, Acetazolamide | 15 | 7.00 | 4.52 |
|  | CWI, No acetazolamide | 103 | 12.16 | 7.17 |
|  | Bariatric surgery | 211 | 8.55 | 6.71 |

CWI: community weight management intervention arm.

A population defined by a PMD between -2dB and -7dB at baseline was identified and analysed to simulate a medically treated population. In this enriched population there was a similar distribution of changes in the point-sensitive deviation at baseline (Figure 2b). Overall, the vast majority of data points that could be analysed were in the 0 to -10Db category (n=4123), compared with points between -10dB and -25dB (n=346) and -25dB to - 5dB, (n=487), respectively. (Table 5)

**Table 5.**
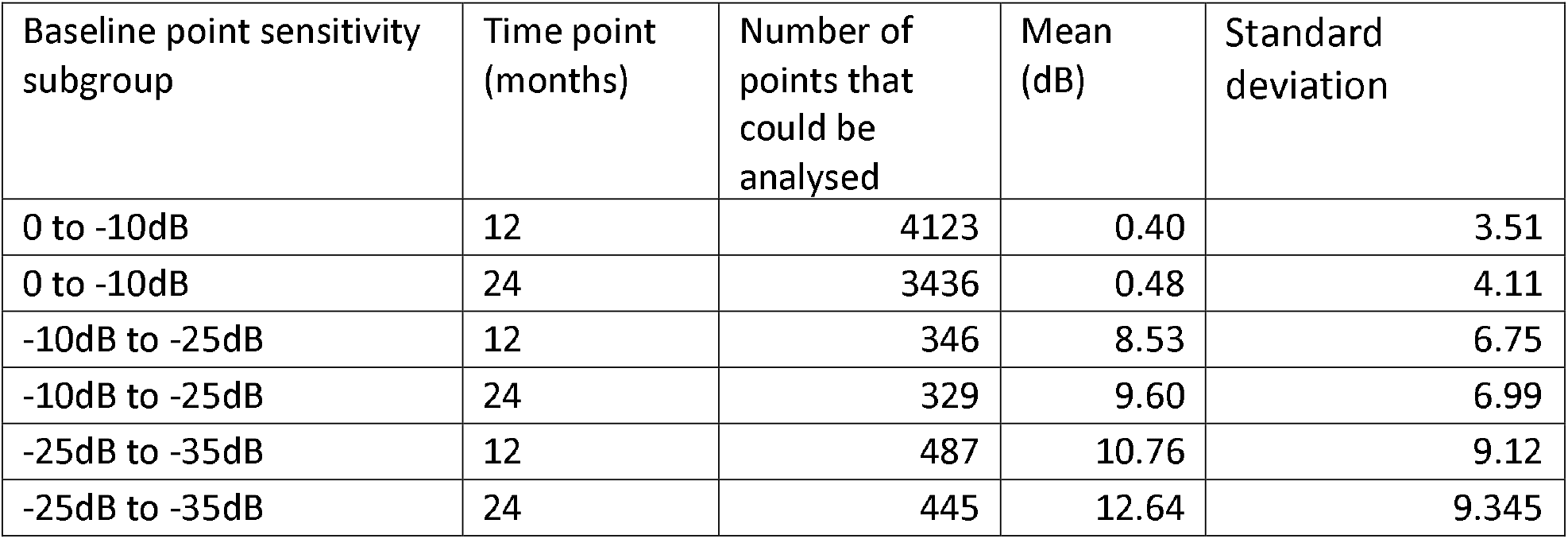
In an enriched population, defined by a PMD between -2dB and -7dB at baseline, the number of point sensitives were categorized by the location point sensitivity.

| Baseline point sensitivity subgroup | Time point (months) | Number of points that could be analysed | Mean (dB) | Standard deviation |
| --- | --- | --- | --- | --- |
| 0 to -10dB | 12 | 4123 | 0.40 | 3.51 |
| 0 to -10dB | 24 | 3436 | 0.48 | 4.11 |
| -10dB to -25dB | 12 | 346 | 8.53 | 6.75 |
| -10dB to -25dB | 24 | 329 | 9.60 | 6.99 |
| -25dB to -35dB | 12 | 487 | 10.76 | 9.12 |
| -25dB to -35dB | 24 | 445 | 12.64 | 9.345 |

### Analysis of pointwise sensitivities in the trial cohort

To establish how point-sensitivity analysis performs in this cohort, we initially looked at the utility of baseline points between -0 and -10dB. As expected, these demonstrated very little change at 12 and 24 months (at 12 months, mean change 0.4 ± 3.5dB; at 24 months, mean change 0.48 ± 4.11 dB). We then selected baseline sensitivities between -10 and -25dB, as we have shown that these have ability to change over time (Figure 2). Indeed, amongst those with baseline -10 to -25dB, it was only when using the whole cohort that the largest mean change was noted: 8.53 ± 6.75dB at 12 months and 9.60 ± 6.99dB at 24 months (Table 4). However, there were fewer points available for analysis n=346 at 12 months and n=329 at 24 months compared with n=4123 at 12 months and n=1844 at 24 months in cases where the baseline pointwise sensitivity ranged from 0 to -10dB. (Table 5) Furthermore, there was little difference between trial arms observed when analysing the points that were between - 10 to -25dB at baseline: bariatric surgery arm was 8.75 ± 6.51 dB at 12 months and CWI was 8.16 ± 7.15dB change at 12 months. This was despite a significant difference between baseline and 12 months in the ICP of -6.0cm CSF between the trial arms. Amongst those in the CWI arm who were not on acetazolamide, point sensitivity mean change between baseline and 12 months was 8.03 ±7.26 dB. Despite the significant reduction in ICP found between the bariatric surgery group as compared to CWI, there was little discrimination analysing point sensitivities between bariatric surgery, CWI, CWI with no acetazolamide treatment, and CWI with concurrent use of acetazolamide. (Table 6)

**Table 6.** Longitudinal mean pointwise location sensitivity changes in those with point sensitivities between -10dB to -25dB, categorized by treatment at 12 and 24 months. CWI, community weight management intervention; SD, standard deviation.

|  | Total cohort |  | Bariatric surgery |  | ALL CWI |  | CWI (no concurrent acetazolamide treatment) |  | CWI and concurrent use of acetazolamide |  |
| --- | --- | --- | --- | --- | --- | --- | --- | --- | --- | --- |
|  | Mean (±SD), number | Effect size | Mean (±SD), number | Effect size | Mean (±SD), number | Effect size | Mean (±SD), number | Effect size | Mean (±SD), number | Effect size |
| 12 months | 8.53 (6.8), 346 | 1.27 | 8.75 (6.5), 219 | 1.34 | 8.16 (7.2), 127 | 1.14 | 8.77 (4.3), 22 | 2.04 | 8.03 (7.6), 105 | 1.05 |
| 24 months | 9.60 (6.99), 329 | 1.37 | 8.55 (6.7), 211 | 1.27 | 11.5 (7.1), 118 | 1.62 | 12.16 (7.2), 103 | 1.69 | 7.00 (4.52), 15 | 1.55 |

CWI, community weight management intervention; SD, standard deviation.

### Categorising the population by baseline PMD

To understand how representative a baseline point sensitivity beyond -10dB in one or more points was in an active IIH population, we calculated the number of points ≥-10dB in each individual (Table 7). In the whole cohort, the median number of points on the baseline visual field worse than -10dB was 5 (IQR 2, 13), with 57% of the cohort having at least 2 points worse than -10dB at baseline (Table 7). The population then was enriched to reflect a potentially medically managed cohort, those with a PMD between -2 and -7dB at baseline. In this group at baseline, 42% had 5 points or more that were worse than -10dB. As the number of points required for analysis decreases, more participants are available for inclusion; for example, 73% had at least 2 or more points and 85% had 1 point worse than - 10dB. (Table 7) 31% of patients at 12 months and 38% at 24 months would have achieved 5 or more points that improved by 7dB. (Table 8)

**Table 7.** Number of participants who had one or more baseline points in either eye with a sensitivity worse than -10dB in the entire cohort with and without perimetric mean deviation criteria. The median points (IQR) column shows the median number of points ≤ -10 in either eye at baseline (and IQR) in only those patients who have at least one qualifying point.

| | | Number of points $\leq -10$dB in either eye at<br>baseline (%) | | | | | |
| --- | --- | --- | --- | --- | --- | --- | --- |
| Population | n | 1 | 2 | 3 | 4 | 5 | Median points<br>(IQR) |
| Whole cohort | 58 | 39<br>(0.672) | 33<br>(0.569) | 26<br>(0.448) | 20<br>(0.345) | 20<br>(0.345) | 5 (2, 13.5) |
| PMD ≥ -2dB | 38 | 33<br>(0.868) | 29<br>(0.763) | 22<br>(0.579) | 19<br>(0.500) | 19<br>(0.500) | 6 (2, 15) |
| Between PMD<br>2dB and 7dB | 33 | 28<br>(0.848) | 24<br>(0.727) | 17<br>(0.515) | 14<br>(0.424) | 14<br>(0.424) | 4 (2, 10) |
| PMD ≤ -7dB | 6 | 6<br>(1.000) | 6<br>(1.000) | 6<br>(1.000) | 6<br>(1.000) | 6<br>(1.000) | 50.5 (32.75,<br>71.25) |

PMD, perimetric mean deviation; IQR, interquartile range

**Table 8.** The percentage and number of participants who had pointwise improvement of 7dB or more from baseline at 12 and 24 months.

| Number of<br>available points<br>on the HVF 24-2<br>at baseline | % number of patients (n) who had a pointwise<br>improvement of 7dB or more from baseline at: |  |
| --- | --- | --- |
|  | 12 months (n=53) | 24 months (n=44) |
| 0 | 26 (14) | 18 (8) |
| 1 | 13 (7) | 9 (4) |
| 2 | 6 (3) | 7 (3) |
| 3 | 4 (2) | 7 (3) |
| 4 | 1 (7) | 1 (4) |
| 5 | 1 (4) | 1 (5) |
| > 5 | 30 (16) | 37 (17) |

## Discussion

We have characterised the pointwise pattern of visual field change in a cohort of people with active IIH who were recruited to the IIH:WT. We observed that those with baseline point sensitivities between 0 and -10dB showed small changes over time and, as expected, were unlikely to demonstrate clinically meaningful change over both 12 and 24 months. Points in the -10 to -25dB category did demonstrate change that could be considered clinically meaningful (mean 8.5dB in at least one point in the whole visual field); however, using data between -10 and -25dB generated fewer data points and large standard deviations for analysis. To emphasize how non-representative a baseline point sensitivity beyond -10dB in one or more points was in our active IIH population, we found that even though the median number of points worse than -10dB was five, 43% of all the participants had fewer than two points worse than -10dB at baseline. Hence, data points worse than - 10dB were not representative of the majority of IIH patients.

Eligibility for the IIH:WT was not determined by PMD criteria. Therefore to enrich the HVF data to reflect a typically medically managed cohort, we chose the baseline HVF in which the PMD was between -2 and -7dB (the range criteria used in the IIHTT^9^). Amongst this group (−2 to -7dB PMD at baseline) 42% had five points or more worse than -10dB at baseline (Table 7). If only two points were required for analysis in the -2 to -7dB population, 73% had two or more points worse than -10dB in either eye at baseline (Table 7). This indicates that the feasibility of using point analysis as an outcome for an interventional medical trial would be challenging, as the pool of point-sensitivity data available for meaningful analysis would be extremely small. Additionally, the participants overall would be less representative of the whole disease spectrum, which may affect the applicability of the results being directly translatable to clinical practice. Lastly, test locations with 8 to 18 dB of loss at baseline had a 95% prediction interval that nearly covered the full measurement range of the instrument (0-40 dB)^28^; this results in the retest variability of these test locations being so poor that there is little signal to be extracted from the variability related noise.^29^

There is no universally adopted minimally clinically important change in any given visual field test parameter in IIH, unlike has been agreed in glaucoma.^30,31,32^ In the IIHTT, effect size was chosen at 1.3dB based on a determination of the change in PMD that would alter patient management by experts. The investigators subsequently concluded that a 0.71dB difference in the PMD between the two trial arms was clinically meaningful.^9,13^ This conclusion was based on significant improvements in lumbar puncture opening pressure, papilloedema (as measured by OCT), and both vision-specific and overall health-related quality of life measures ^9,13^

Most of the literature analysing HVF data is focused on glaucoma, the most common optic neuropathy.^33,34^ In glaucoma, visual field progression equal to or faster than ™0.5 dB per year for at least five abnormal test locations at baseline has been found to be clinically significant,^35^ as have changes from baseline beyond the 5% probability levels for the Glaucoma Change Probability analysis, in five or more reproducible points, or visual field locations.^31^ Pointwise linear regression has been demonstrated to be a sensitive technique for detecting rate of progressive deterioration in visual fields in glaucoma.36 The IIHTT investigators found that pointwise visual field change improvements were identified around the blind spot and the nasal area, possibly reflecting the reduction in optic head nerve swelling as papilloedema resolved.^14^ Overall, there are fundamental differences between the two diseases, which could potentially confound the applicability of glaucoma outcome measures to IIH trials. Other tools that assess visual function, such visual acuity (Snellen or LogMar), colour vision, and contrast sensitivity, have not been found to be discriminatory in medically managed IIH.^9,37^ The IIH patient experience, as measured by specific quality-of-life tools, was found to be influenced mainly by PMD in the better eye, visual acuity in the worse eye, visual symptoms, and pain symptoms in the IIHTT cohort.^38^ Other studies analysing global quality of life tools (SF-36) have identified headache as a major factor.^39,40^ Indeed, in one study, whilst both headache and vision featured in the top 10 priorities, headache ranked higher than vision.^41^

One key limitation of this study is that it only included patients in a well-established disease cohort. Hence, the results may not be applicable to those with recently diagnosed IIH, as were recruited to the IIHTT,^9^ and may not be applicable to severely affected patients that may require emergency surgery.^6,42^ Regression to the mean was demonstrated in our cohort for the mean deviation (Figure 2), and although this occurs in many diseases over time, it provides evidence that a control arm is fundamental to the design of interventional trials in IIH.

As we demonstrated in this study and as has been reported by others^14^ the visual field deficit in IIH typically occurs across the full VF and increases with eccentricity.^17^ Unfortunately, these are the very points that show the largest variability in visual field testing.^43,44^ Visual field tests also have been found to be unreliable when visual field locations have sensitivity below 15 to 19 dB because of a reduction in the asymptotic maximum response probability.^45^ In addition to these limitations of the test, there are demonstrable changes in cognition in the domains of attention and executive function that have been found in IIH patients and that directly affect the performance indicators in visual field testing.^12^

These data suggest that although point analysis of the HVF in active, medically treated IIH is possible, this technique is considerably compromised as the amount of data available to demonstrate a clinically meaningful change is small. Hence, point analysis seems to offer no advantage over analysis of global PMD scores in this population. Future studies may consider investigating the use of a larger stimulus size which has been demonstrated to retain the ability to detect defects, lower retest variability and improve the useful dynamic range of the instrument.^46,47^ Recently, archetypal analysis, an unsupervised machine-learning technique, has been reported in IIH. Archetypal analysis could help reduce dependence on subjective clinician visual field interpretation and could improve the accuracy and uniformity of visual field analysis by monitoring regional defects.^48^ In the IIHTT, the investigators identified a subtle, mild general HVF depression pattern in the participants at baseline. This is a notable finding, as visual fields with global diffuse damage tend to be more variable than fields with focal blinding damage and other regions that are normal,^49^ further highlighting that more research on the utility of automated visual fields in IIH is required.

In this study of patients with active IIH, pointwise sensitivity analysis did not confer an advantage over global PMD. As expected, baseline points that were better than -10dB had little room to improve over time and, thus, offered little utility for analysis. If the generic threshold for a clinically meaningful change of 7dB, as suggested by the National Eye Institute (NEI) of the National Institutes of Health (NIH) for glaucoma treatment trials, is required in IIH treatment trials, baseline points in the range of -10 to -25dB would be needed for analysis. However, this approach appears not to be feasible in IIH as in our study there were too few data points available for analysis, and these points are known to be more variable as they become more negative. Even when a limited threshold was set to determine a clinically meaningful change, point analysis did not appear to offer an advantage over global PMD. Consequently, point sensitivity analysis in medically treated IIH is likely to be prohibitive to running clinical trials and unrepresentative of the disease spectrum seen in IIH.

## Data Availability

Anonymised individual participant data will be made available along with the trial protocol and statistical analysis plan. Proposals should be made to the corresponding author and will be reviewed by the Birmingham Clinical Trials Unit Data Sharing Committee in discussion with the Chief Investigator. A formal Data Sharing Agreement may be required between respective organisations once release of the data is approved and before data can be released.

## Acknowledgements

None

---

**Supplementary Table 1.** IIH symptoms at diagnosis, as reported by participants following direct questioning

| Symptoms reported | Total n=66<br>(%) |
| --- | --- |
| Headache |  |
| Yes | 63 (95) |
| No | 3 (5) |
| Visual symptoms* |  |
| Yes | 48 (73) |
| No | 18 (27) |
| Diplopia |  |
| Yes | 18 (27) |
| No | 48 (73) |
| Visual Obscurations |  |
| Yes | 34 (52) |
| No | 32 (52) |
| Pulsatile Tinnitus |  |
| Yes | 49 (74) |
| No | 17 (26) |

\*Visual symptoms, as defined by the patient, that did not include diplopia or visual
obscurations such a blurred vision or difficulty reading.

## References

1. Mollan SP, Grech O, Alimajstorovic Z, Wakerley BR, Sinclair AJ. New horizons for idiopathic intracranial hypertension: advances and challenges. Br Med Bull. 2020 Dec 15;136(1):118–126

2. Westgate CS, Botfield HF, Alimajstorovic Z, Yiangou A, Walsh M, Smith G, Singhal R, Mitchell JL, Grech O, Markey KA, Hebenstreit D, Tennant DA, Tomlinson JW, Mollan SP, Ludwig C, Akerman I, Lavery GG, Sinclair AJ. Systemic and adipocyte transcriptional and metabolic dysregulation in idiopathic intracranial hypertension. JCI Insight. 2021 May 24;6(10):e145346.

3. O’Reilly MW, Westgate CS, Hornby C, Botfield H, Taylor AE, Markey K, Mitchell JL, Scotton WJ, Mollan SP, Yiangou A, Jenkinson C, Gilligan LC, Sherlock M, Gibney J, Tomlinson JW, Lavery GG, Hodson DJ, Arlt W, Sinclair AJ. A unique androgen excess signature in idiopathic intracranial hypertension is linked to cerebrospinal fluid dynamics. JCI Insight. 2019 Mar 21;4(6):e125348.

4. Mollan SP, Tahrani AA, Sinclair AJ. The Potentially Modifiable Risk Factor in Idiopathic Intracranial Hypertension: Body Weight. Neurol Clin Pract. 2021 Aug;11(4):e504–e507

5. Adderley NJ, Subramanian A, Nirantharakumar K, Yiangou A, Gokhale KM, Mollan SP, Sinclair AJ. Association Between Idiopathic Intracranial Hypertension and Risk of Cardiovascular Diseases in Women in the United Kingdom. JAMA Neurol. 2019 Sep 1;76(9):1088–1098.

6. Mollan SP, Mytton J, Tsermoulas G, Sinclair AJ. Idiopathic Intracranial Hypertension: Evaluation of Admissions and Emergency Readmissions through the Hospital Episode Statistic Dataset between 2002-2020. Life (Basel). 2021;11(5):417

7. Mollan SP, Mitchell JL, Ottridge RS, Aguiar M, Yiangou A, Alimajstorovic Z, Cartwright DM, Grech O, Lavery GG, Westgate CSJ, Vijay V, Scotton W, Wakerley BR, Matthews TD, Ansons A, Hickman SJ, Benzimra J, Rick C, Singhal R, Tahrani AA, Brock K, Frew E, Sinclair AJ. Effectiveness of Bariatric Surgery vs Community Weight Management Intervention for the Treatment of Idiopathic Intracranial Hypertension: A Randomized Clinical Trial. JAMA Neurol. 2021 Jun 1;78(6):678–686.

8. Mollan SP, Sinclair AJ. Outcomes measures in idiopathic intracranial hypertension. Expert Rev Neurother. 2021 Jun;21(6):687–700.

9. NORDIC Idiopathic Intracranial Hypertension Study Group Writing Committee, Wall M, McDermott MP, Kieburtz KD, Corbett JJ, Feldon SE, Friedman DI, Katz DM, Keltner JL, Schron EB, Kupersmith MJ. Effect of acetazolamide on visual function in patients with idiopathic intracranial hypertension and mild visual loss: the idiopathic intracranial hypertension treatment trial. JAMA. 2014;311(16):1641–51

10. Cello KE, Keltner JL, Johnson CA, Wall M; NORDIC Idiopathic Intracranial Hypertension Study Group. Factors Affecting Visual Field Outcomes in the Idiopathic Intracranial Hypertension Treatment Trial. J Neuroophthalmol. 2016 Mar;36(1):6–12

11. Junoy Montolio FG, Wesselink C, Gordijn M, Jansonius NM. Factors that influence standard automated perimetry test results in glaucoma: test reliability, technician experience, time of day, and season. Invest Ophthalmol Vis Sci. 2012 Oct 9;53(11):7010–7

12. Grech O, Coulter A, Mitchell JL, et al. Cognitive performance in idiopathic intracranial hypertension and relevance of intracranial pressure, Brain Communications, 2021;, fcab202, https://doi.org/10.1093/braincomms/fcab202

13. Bruce BB, Digre KB, McDermott MP, Schron EB, Wall M; NORDIC Idiopathic Intracranial Hypertension Study Group. Quality of life at 6 months in the Idiopathic Intracranial Hypertension Treatment Trial. Neurology. 2016;87(18):1871–1877.

14. Wall M, Johnson CA, Cello KE, Zamba KD, McDermott MP, Keltner JL; NORDIC Idiopathic Intracranial Hypertension Study Group. Visual Field Outcomes for the Idiopathic Intracranial Hypertension Treatment Trial (IIHTT). Invest Ophthalmol Vis Sci. 2016;57(3):805–812

15. Heijl A, Lindgren G, Olsson J. A package for the statistical analysis of visual fields. Doc Ophthalmol Proc Ser. 1987;49:153–168

16. Fitzke FW, Hitchings RA, Poinoosawmy DP, McNaught AI, Crabb DP. Analysis of visual field progression in glaucoma. Br J Ophthalmol. 1996;80:40–48

17. Wall M, Subramani A, Chong LX, Galindo R, Turpin A, Kardon RH, Thurtell MJ, Bailey JA, Marin-Franch I. Threshold Static Automated Perimetry of the Full Visual Field in Idiopathic Intracranial Hypertension. Invest Ophthalmol Vis Sci. 2019 May 1;60(6):1898–1905

18. Heijl A Lindgren G Olsson J. Normal variability of static perimetric threshold values across the central visual field. Arch Ophthalmol. 1987; 105: 1544–1549

19. Heijl A Asman P. A clinical study of perimetric probability maps. Arch Ophthalmol. 1989;107: 199–203.

20. Wall M, Doyle CK, Zamba KD, Artes P, Johnson CA. The repeatability of mean defect with size III and size V standard automated perimetry. Invest Ophthalmol Vis Sci. 2013 Feb 15;54(2):1345–51

21. Ottridge R, Mollan SP, Botfield H, Frew E, Ives NJ, Matthews T, Mitchell J, Rick C, Singhal R, Woolley R, Sinclair AJ. Randomised controlled trial of bariatric surgery versus a community weight loss programme for the sustained treatment of idiopathic intracranial hypertension: the Idiopathic Intracranial Hypertension Weight Trial (IIH:WT) protocol. BMJ Open. 2017 Sep 27;7(9):e017426

22. Friedman DI, Liu GT, Digre KB. Revised diagnostic criteria for the pseudotumor cerebri syndrome in adults and children. Neurology. 2013 Sep 24;81(13):1159–65

23. Frisén L. Swelling of the optic nerve head: a staging scheme. J Neurol Neurosurg Psychiatry. 1982;45(1):13–18

24. Yohannan J, Wang J, Brown J, Chauhan BC, Boland MV, Friedman DS, Ramulu PY. Evidence-based Criteria for Assessment of Visual Field Reliability. Ophthalmology. 2017 Nov;124(11):1612–1620.

25. Van Rossum, G. & Drake Jr, F.L., 1995. Python reference manual, Centrum voor Wiskunde en Informatica Amsterdam

26. Saifee, M., Wu, J., Liu, Y., Ma, P., Patlidanon, J., Yu, Y., Ying, G. and Han, Y., 2021. Development and Validation of Automated Visual Field Report Extraction Platform Using Computer Vision Tools. Frontiers in Medicine, 8.

27. opensource.google. 2021. Projects – opensource.google. [online] Available at: <https://opensource.google/projects/tesseract> [Last accessed 6 October 2021].

28. Heijl A, Lindgren A, Lindgren G. Test-retest variability in glaucomatous visual fields. Am J Ophthalmol. 1989;108(2):130–135.

29. Wall M, Artes P, Zamba KD. The Effective Dynamic Ranges for Glaucomatous Visual Field Progression with Standard Automated Perimetry and Stimulus Sizes III and V. Invest Ophthalmol Vis Sci 2018;59:439–445 https://doi.org/10.1167/iovs.17-22390.

30. Csaky KG, Richman EA, Ferris FL 3rd. Report from the NEI/FDA Ophthalmic Clinical Trial Design and Endpoints Symposium. Invest Ophthalmol Vis Sci. 2008 Feb;49(2):479–89

31. Weinreb RN, Kaufman PL. The glaucoma research community and FDA look to the future: a report from the NEI/FDA CDER Glaucoma clinical trial design and endpoints symposium. Invest Ophthalmol Vis Sci. 2009;50:1497–1505

32. Heijl A, Leske MC, Bengtsson B, Bengtsson B, Hussein M; Early Manifest Glaucoma Trial Group. Measuring visual field progression in the Early Manifest Glaucoma Trial. Acta Ophthalmol Scand. 2003;81(3):286–293.

33. Bourne RRA, Taylor HR, Flaxman SR, et al. Vision Loss Expert Group of the Global Burden of Disease Study. Number of people blind or visually impaired by glaucoma worldwide and in world regions 1990-2010: a meta-analysis. PLoS One. 2016;11(10):e0162229

34. Musch DC, Gillespie BW, Lichter PR, Niziol LM, Janz NK; CIGTS Study Investigators. Visual field progression in the Collaborative Initial Glaucoma Treatment Study: the impact of treatment and other baseline factors. Ophthalmology. 2009;116(2):200–207.

35. De Moraes CG, Liebmann JM, Levin LA. Detection and measurement of clinically meaningful visual field progression in clinical trials for glaucoma. Prog Retin Eye Res. 2017 Jan;56:107–147

36. Gardiner SK, Crabb DP. Examination of Different Pointwise Linear Regression Methods for Determining Visual Field Progression. Invest. Ophthalmol. Vis. Sci.2002;43(5):1400–1407

37. Sinclair AJ, Burdon MA, Nightingale PG, Ball AK, Good P, Matthews TD, Jacks A, Lawden M, Clarke CE, Stewart PM, Walker EA, Tomlinson JW, Rauz S. Low energy diet and intracranial pressure in women with idiopathic intracranial hypertension: prospective cohort study. BMJ. 2010 Jul 7;341:c2701

38. Digre KB, Bruce BB, McDermott MP, Galetta KM, Balcer LJ, Wall M; NORDIC Idiopathic Intracranial Hypertension Study Group. Quality of life in idiopathic intracranial hypertension at diagnosis: IIH Treatment Trial results. Neurology. 2015 Jun 16;84(24):2449–56

39. Mulla Y, Markey KA, Woolley RL, Patel S, Mollan SP, Sinclair AJ. Headache determines quality of life in idiopathic intracranial hypertension. J Headache Pain. 2015;16:521

40. Kleinschmidt JJ, Digre KB, Hanover R. Idiopathic intracranial hypertension: relationship to depression, anxiety, and quality of life. Neurology. 2000 Jan 25;54(2):319–24

41. Mollan S, Hemmings K, Herd CP, Denton A, Williamson S, Sinclair AJ. What are the research priorities for idiopathic intracranial hypertension? A priority setting partnership between patients and healthcare professionals. BMJ Open. 2019 Mar 15;9(3):e026573

42. Mollan SP, Davies B, Silver NC, Shaw S, Mallucci CL, Wakerley BR, Krishnan A, Chavda SV, Ramalingam S, Edwards J, Hemmings K, Williamson M, Burdon MA, Hassan-Smith G, Digre K, Liu GT, Jensen RH, Sinclair AJ. Idiopathic intracranial hypertension: consensus guidelines on management. J Neurol Neurosurg Psychiatry. 2018 Oct;89(10):1088–1100

43. Keltner JL, Johnson CA, Quigg JM, Cello KE, Kass MA, Gordon MO. Confirmation of visual field abnormalities in the Ocular Hypertension Treatment Study. Ocular Hypertension Treatment Study Group. Arch Ophthalmol. 2000 Sep;118(9):1187–94

44. Wall M, Brito CF, Woodward KR, Doyle CK, Kardon RH, Johnson CA. Total Deviation Probability Plots for Stimulus Size V Perimetry: A Comparison with Size III Stimuli. Arch Ophthalmol 2008;126:473–479; doi: 10.1001/archopht.126.4.473.PMID:18413515.

45. Gardiner SK, Swanson WH, Goren D, Mansberger SL, Demirel S. Assessment of the reliability of standard automated perimetry in regions of glaucomatous damage. Ophthalmology. 2014 Jul;121(7):1359–69

46. Wall M, Artes P, Zamba KD. The Effective Dynamic Ranges for Glaucomatous Visual Field Progression with Standard Automated Perimetry and Stimulus Sizes III and V. Invest Ophthalmol Vis Sci 2018;59:439–445.

47. Gardiner SK. Differences in the Relation Between Perimetric Sensitivity and Variability Between Locations Across the Visual Field. Invest Ophthalmol Vis Sci. 2018 Jul 2;59(8):3667–3674.

48. Doshi H, Solli E, Elze T, Pasquale LR, Wall M, Kupersmith MJ. Unsupervised Machine Learning Identifies Quantifiable Patterns of Visual Field Loss in Idiopathic Intracranial Hypertension. Transl Vis Sci Technol. 2021 Aug 2;10(9):37.

49. Russell RA, Garway-Heath DF, Crabb DP. New insights into measurement variability in glaucomatous visual fields from computer modelling. PLoS One. 2013 Dec 30;8(12):e83595.

